# Effect of the Gut Microbiome on the reduction of uremic toxins in patients with chronic kidney disease: A systematic review & Network meta-analysis

**DOI:** 10.1101/2025.02.21.25322662

**Authors:** Renata Cedillo-Flores, Miguel Angel Cuevas-Budhart, Iván Cavero Redondo, Maria Soledad Kappes, Marcela Ávila, Ramón Paniagua

## Abstract

**Introduction:** Chronic kidney disease (CKD) is associated with increased intestinal barrier permeability, leading to heightened inflammation and oxidative stress. These changes contribute to complications such as cardiovascular disease, anemia, altered mineral metabolism, and CKD progression. Interventions using prebiotics, probiotics, and synbiotics may help mitigate dysbiosis and improve intestinal barrier function.

**Aim:** This study conducted a network meta-analysis to evaluate the effectiveness of probiotics, prebiotics, and synbiotics in reducing uremic toxins produced by the gut microbiota in CKD patients.

**Method:** A systematic review and network meta-analysis of randomized clinical trials (RCTs) was performed. The analysis focused on the use of prebiotics, probiotics, and synbiotics in CKD patients at stages 3 to 5, as per KDIGO guidelines, and their association with reductions in uremic toxins such as indoxyl sulfate (IS), p-cresyl sulfate (pCS), urea, and creatinine. The study follows the PRISMA statement.

**Results:** The studies included 331 patients, primarily male, across CKD stages 3a to 5. The interventions showed a positive impact on the gut microbiota composition, leading to reductions in both free and total p-cresyl sulfate and indoxyl sulfate.

**Conclusion:** The findings suggest that modulating the gut microbiota through these interventions can effectively reduce specific uremic toxins. However, further trials are necessary to better understand the microbiota modulation and its impact on intestinal bacterial composition.

**PROSPERO Registration:** CRD42023438901

## 1. INTRODUCTION

Various conditions present in patients with chronic kidney disease (CKD) are associated with the development of alterations in the permeability of the intestinal barrier and the Gut microbiome (GM), among which are the loss of renal function, uremic toxicity, and the use of frequent antibiotics(1).

Changes in the quantitative and qualitative composition of the intestinal microbial population have been implicated in the pathogenesis of different diseases, including the systemic inflammatory state, CKD progression, and CKD-related cardiovascular complications; this is highly dependent on substantial changes, for example, in diet composition and nutrient intake due to recommended restrictions to prevent the aforementioned complications(2,3)

It is important to add that the GM changes constantly during the course of CKD, which produces a metabolic load that could further increase cardiovascular risk. In addition, metabolites derived from the gut microbiota, including products of protein or choline fermentation, such as p-cresyl sulfate (PCS) and indoxyl sulfate (IS), may contribute to decreased renal function and worsen kidney function and cardiovascular disease(2).

### 1.1 Probiotics, prebiotics and symbiotics

Probiotics are live microorganisms that, when administered in adequate amounts, confer a health benefit on the host(4), (5). On the other hand, prebiotics are mostly fibers that are indigestible food ingredients and beneficially affect host health by selectively stimulating the growth or activity of some genera of microorganisms in the colon, *typically lactobacilli* and *bifidobacteria. Lastly, synbiotics* are next-generation probiotics made with various formulations of probiotics and prebiotics that work synergistically to restore healthy gut ecology (6–8).

In this sense, the term immunonutrition can be defined as the area of nutrition dedicated to the study of the processes by which nutrients modulate the actions of the immune system and their use for this purpose. This encompasses factors related to nutrition, infection, inflammation, tissue damage, and immunity, with a key role in the immune, endocrine, nervous, and microbiota systems.(9,10).

### 1.2 Uremic toxins, CKD and the risk of cardiovascular disease

P-Cresol can be considered a prototypical protein-bound uremic toxin, and indoxyl sulfates a circulating uremic toxin. Both are excreted via the kidneys; therefore, serum concentrations increase progressively as GFR decreases. These metabolites cause vascular endothelial cell injury by increasing leukocyte activation and adhesion and contributing to local inflammation and oxidative stress(11,12).

Furthermore, higher concentrations of circulating IS and PCS correlate with measures of vascular dysfunction and aortic calcification and are independently associated with cardiovascular disease among people with kidney disease (11,12).

The GM is altered in CKD patients due to increased intestinal permeability and the accumulation of uremic toxins in the plasma (causing vascular alterations). Various therapeutic interventions have recently been explored to improve intestinal microbiota dysbiosis, such as the use of prebiotics, probiotics, and symbiotics. These could reduce the generation of uremic toxins by increasing or decreasing the associated bacteria, reducing the cardiovascular effects that CKD brings with it(13).

Some related studies in this context are Pan et all (14), who found a decrease in C-reactive protein (CRP) and IL-6 levels after two months of treatment in a group of 58 patients treated with probiotics; in addition, higher levels were obtained in the domains of physical functioning and social functioning than the control group.

In another study, Raquel Armani et al(15), found that the prebiotic FOS reduced circulating levels of IL-6 in patients with CKD and preserved endothelial function only in those with less damaged endothelium. No effect of FOS on arterial stiffness was observed.

This work aims to analyze the relationship between probiotics, prebiotics, and symbiotics in the reduction of uremic toxins produced by the intestinal microbiota.

## 2. METHODOLOGY

### 2.1 Design

A systematic review and Network meta-analysis of randomized clinical trials (RCTs) was carried out. A systematic and rigorous process was followed for the identification and evaluation of the existing scientific evidence on the relationship between the use of prebiotics, prebiotics, and synbiotics in patients with chronic kidney disease in stages 3 to 5 according to the KDIGO guidelines and associate the use of them with the reduction of uremic toxins: Indoxyl sulfate (IS), p-Cresyl Sulfate (pCS), Urea, Creatinine (Cr), and Phosphate (PHOS).

The recommendations of the PRISMA statement and the Cochrane Manual for systematic reviews and meta-analyses of intervention studies were followed. The process to follow for the development of the investigation included: Formulation of the question, establishment of selection criteria, systematic search, and evaluation of the data through analysis and narrative and integrative synthesis (16,17).

First, the PICO strategy was used to develop the research question: What is the relationship between use of probiotics, prebiotics, and symbiotics in reducing uremic toxins produced by the intestinal microbiota?

### 2.2 Selection criteria

The studies included were randomized controlled trials, published between 2019 and 2023, available in open access and published in English and Spanish. All studies that were not related to the topic were eliminated.

### 2.3 Search strategy

An extensive search was performed in the following databases: Web of Science, Scopus, the Cochrane Register of Controlled Trials, and PubMed. Clinical trials of the population with chronic kidney disease and who received prebiotics, probiotics or oral symbiotic were searched for and selected.

We used the Health Sciences Descriptors (DeCS) and the Medical Subject Headings (MeSH) for the keywords: gut microbiota, gut microbiome, chronic kidney disease, cardiovascular disease, and clinical trial. The Boolean operators used were the intersection (AND) to establish the logical operations between concepts and (OR) to retrieve documents where at least one of the specified arguments appears.

### 2.4 Study selection

A s shown in Figure 1 PRISMA Diagram summarizes the selection process. After eliminating duplicates 1030 articles were selected; the title and abstract were individually inspected, selecting those documents that could be relevant to this study, 952 studies were eliminated. (See Figure 1).

**Figure 1.**
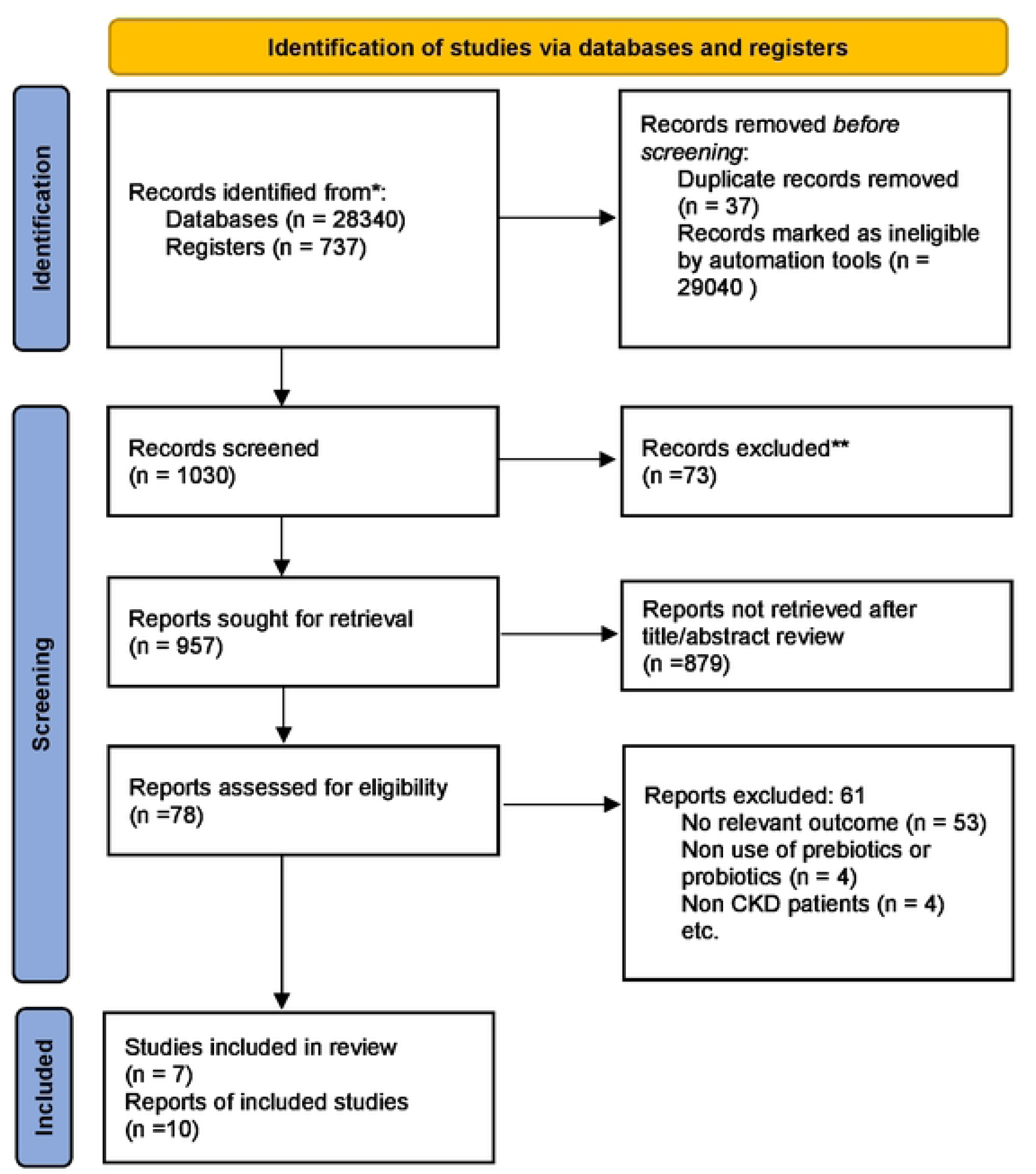
PRISMA 2020 flow diagram for new systematic reviews.

Subsequently, the analysis of the 78 full-text studies was carried out, 68 articles were excluded, mainly because there were no randomized controlled trials, those pilot trials, or study protocols, and they did not adhere to the indicated theme. Probiotics, prebiotics, and symbiotic were included regardless of dose or duration of treatment. Finally, the remaining articles were read in their entirety, and those that were conclusive in their results were chosen.

Any disagreement about the eligibility of certain studexcluies was resolved by a third independent reviewer.

### 2.5 Assessment of Methodological Quality and Risk of Bias

In the selection process, the methodological quality of the included studies was assessed after their selection according to the JADAD criteria (18). This scale evaluates the CS from 0 to 5, depending on the randomization and its method, the type of blinding and its method, and the losses and withdrawals from the study, considering low methodological quality those with a score of less than 3.

To assess the risk of bias, version 2 of the Cochrane risk of bias tool for randomized trials (RoB 2)(19) was used, it is structured on a fixed set of bias dimensions, focusing on different aspects of the design, such as the conduct and report of the test. Within each domain, a series of questions (’flag questions’) aim to elicit information about trial characteristics that are relevant to the risk of bias. An algorithm generates a proposed judgment on the risk of bias arising from each domain, based on the responses to the flagging questions. The judgment may be ‘low’ or ‘high’ risk of bias, or may express ‘some concerns’(20).

For the network analysis, the global method was applied to jointly investigate the presence of inconsistencies in the possible investigated sources in the entire simultaneous network, using a design-by-treatment interaction model(21,22). The reduction of uremic toxins was estimated by strata of probiotics, symbiotics, and prebiotics. The presence of minor study effects for each outcome was assessed by drawing comparison-adjusted funnel plots that fit the different comparisons of immunonutrition use that are included in the study. In addition, a network meta-analysis was performed using the network data package 20 and Network graphs in STATA version 16.0 (StataCorp, College Station, TX, USA).

As study designs and outcome definitions varied, the random effect was used to pool estimates from studies. Pooled estimates were transformed and presented with means and standard deviations. As a sensitivity analysis, standardized residuals were calculated, and outliers with p > 0.05 (based on the t-distribution) were removed. Meta-regression and Odds-Ratios (OR) with 95% confidence intervals (CI) were used.

Additionally, we estimated the surface under the cumulative ranking (SUCRA) for each intervention. SUCRA involves the assignment of a numerical value between 0 and 100 to simplify the classification of each intervention in the rankogram. The best intervention obtained a SUCRA value closest to 100, and the worst intervention obtained a value closest to 0(23).

## 3. RESULTS

### 3.1 Data Availability and Study Characteristics

After the pertinent screenings as well as the scrutiny of the articles in-depth and which met the criteria for inclusion, 13 documents were selected to be assessed for methodological quality and risk of bias with ROB2. Only seven articles were chosen to perform the statistical analysis; the other six included patients that were in renal replacement treatment: four were on HD, and two were on PD, which are added as supplementary material.

Table 1 shows that the selected studies found Italy(15,24,25) as the principal country with more clinical trials in CKD patients, followed by Brazil(26,27). The interventions carried out were three with prebiotics (26–28), two with probiotics (15,24), and two with symbiotics (25,29).

**Table 1.** Characteristics of the included studies.

| Author (year)<br>Country | Aim | Design | Sample<br>size | Intervention | Statistics analysis | Main Results |
| --- | --- | --- | --- | --- | --- | --- |
| De Mauri<br>(2022)<br>Italy(24) | Evaluate whether the association of selected probiotics on top of a low protein diet could reduce the burden of uremic, microbiota-derived, and proatherogenic toxins in patients with advanced renal failure who were not on dialysis. | Placebo-controlled, randomized study | IG= 24<br>CG= 23 | Probiotic | Analysis of blood and urine samples.<br>Shapiro–Wilk and q–q plot tests, Mann–Whitney U-test, Wilcoxon signed rank test, and Kaplan–Meier analysis | Patients in the placebo group showed increased serum values of total cholesterol, LDL cholesterol, lipoprotein-associated phospholipase, and indoxyl-sulphate, while the 24 subjects in the probiotics group showed a trend in reducing microbiota toxins. A reduction of antihypertensive and diuretic medications was possible in the probiotics group. |
| Cosola (2021)<br>Italy(25) | Investigate the effects of the symbiotic on the barrier permeability of different gastrointestinal tracts and on gastrointestinal symptoms assessed by the GSRS questionnaire. | Randomized trial | IG= 23<br>CG= 24 | Symbiotic | Urine and blood samples<br>Linear regression analysis. Mann-Whitney test, Kruskal-Wallis multiple-comparison, Spearman, Fisher’s LSD. | Two-month administration of the symbiotic resulted in a decrease of free IS in the CKD group. After supplementation, reduction of small intestinal permeability and amelioration of abdominal pain and constipation syndromes were observed only in the CKD group. |
| McFarlane<br>(2021)<br>Australia (29) | Evaluate the feasibility of a trial of long-term symbiotic supplementation in adults with stage 3–4 CKD. | Double-blind, placebo-controlled, randomized trial | IG= 28<br>CG= 28 | Symbiotic | Blood and stool samples were collected.<br>Chi-square, Fisher’s exact tests, Student’s t-test, and Mann–Whitney test | No differences between free and total uremic toxins between placebo and symbiotic groups were observed. Symbiotic supplementation resulted in a 3.14 mL/min/1.73 m2 reduction in eGFR and a 20.8 µmol/L increase in serum creatinine concentration. |
| Ramos (2019)<br>Brazil (27) | Investigate the effect of a prebiotic fructooligosaccharide (FOS) on uremic toxins of non-dialysis-dependent CKD (NDD-CKD) patients. | Randomized trial | IG = 23<br>CG n= 23 | Prebiotic | Blood samples were collected.<br>Shapiro–Wilk test, chi-square, Fisher’s exact test, Student’s t-test, and Mann–Whitney test. | There was a trend in the difference between the groups of serum total DPCS and serum-free Δ%PCS. Aside from the decreased high-density lipoprotein cholesterol in the intervention, no differences were observed in the change of IS, IAA, or other secondary outcomes between the groups. |
| Simeoni<br>(2019) Italy<br>(37) | Describe gut dysbiosis in initial stages of CKD. | Clinical trial | IG = 24<br>CG = 23 | Probiotic | Blood, urine, and stool samples were collected.<br>Student’s paired t test, Wilcoxon test, simple t test, Mann-Whitney U test, Pearson. | Mean fecal Lactobacillales and Bifidobacteria concentrations were increased, only in the probiotics group. Conversely, mean urinary indican and 3-MI levels were only in the group treated with probiotics. Compared to the placebo group, significant improvements in C-reactive protein, iron, ferritin, transferrin saturation, β2-microglobulin, serum iPTH, and serum calcium were observed only in the probiotics group. |
| Ebrahim (2022) South Africa (28) | Investigate the effect of a β-glucan prebiotic on kidney function, uremic toxins, and the gut microbiome in stage 3 to 5 CKD participants | Randomized Control Trial | IG = 23<br>CG = 22 | Prebiotic | Blood samples for quantification of uremic toxins and stool samples for characterization of the gut microbiome were obtained.<br>Kolmogorov, Mann–Whitney and chi-square tests. | There was a significant reduction in uremic toxin levels at different time points, in free IxS at 8 weeks and 14 weeks, free pCS at 14 weeks and total and free pCG. There were no differences in relative abundances of gender between groups. The β-glucan prebiotic significantly altered uremic toxin levels of intestinal origin and favorably affected the gut microbiome. |
| Armani (2019) Brazil (26) | Evaluate the effect of the prebiotic FOS on endothelial function and arterial stiffness in non-dialysis CKD patients | Randomized controlled trial | IG = 23<br>CG = 23 | Prebiotic | Fasting blood samples were collected.<br>Kolmogorov–Smirnov, g Student’s t-test, Mann–Whitney U-test, Wilcoxon, chi-squared analysis, Fischer’s and McNemer. | There was a significant decrease in IL-6 levels and a trend toward PCS reduction only in the prebiotic group. Comparing both groups, there was no difference in FMD and PWV. FMD remained stable in the prebiotic group and decreased in the placebo group. |

CKD: chronic kidney disease, PCS: p-cresyl sulfate; IxS: indoxyl sulfate; IAA: indole-3-acetic acid; LPS: lipopolysaccharide; FOS: fructooligosaccharide; IL-6: interleukin-6; iPTH: intact parathyroid hormone; FMD: flow-mediated dilatation; PWV: pulse wave velocity

The included studies were mostly single-center (15,24–28), and only one with two tertiary renal care outpatient departments. The total sample was 331 patients. Regarding the gender of the participants, there was a higher prevalence of men compared to women.

All included only adults with the KDOQI classification who did not participate in dialysis treatment. The patients enrolled in the studies were in stages from 3a to 5, of which only one was performed with stage 3a patients(15) and one from stage IV to stage V(24).

Regarding the interventions, Ramos et al. (27) and Armani et al. (26) used prebiotics in sachet presentation with 12 g each, and Ebrahim et al. (28) used a dose of 13.5 g. The intake was after food (lunch or dinner) since they dissolved in water; Armani et al. (28) was two times a day (6g each) and also recommended using it added to food (smoothies, cereal, yogurt, etc.). It should be emphasized that Ramos et al (27) progressively raised the dose of prebiotics to avoid gastrointestinal symptoms, starting with three grams, and increasing by three every three days until reaching 12g.

McFarlane et al. (29) and Cosola et al. (30) used symbiotics. The first consisted of 20g/day of a high-resistant starch fiber supplement and the probiotic component taken in the morning with food, and the second of 2 bags/day of a mixture of probiotics, prebiotics, and natural antioxidants. McFarlane also had a two-week dose escalation in which the daily dose was 10g and then 20g.

Lastly, the use of probiotics was carried out by De Mauri et al. (31) and Simeoni et al. (15); the first used strains of *Bifidobacterium and Lactobacillus*, administered two doses per day for one month and one dose for the following two months. In the case of Simeoni, the treated group underwent three phases of treatment using different probiotics: intestinal cleaning, intestinal colonization, and microbiota maintenance. Phase 1 used one capsule during main meals for one week of a complex of probiotics. The intestinal colonization lasted for two weeks of one capsule of a complex of *Bifidobacteria* and one capsule of *Lactobacillus* once a day; and phase 3 used both *Bisifelle* and *Ramnoselle* oral administration, one capsule of each, twice per day during breakfast and dinner for three months.

The follow-up period of the studies to evaluate the effect of the interventions was two months for the shortest(25), followed by two and a half months(28), three months(26,27), one year(29), and the longest of three years(24). During the follow-up, the patients were evaluated at the beginning of the study, during the treatment and at the end of it, except with Simeoni, which only evaluate the beginning and the end.

Regarding biochemical parameters that had changes exclusively in the intervention group, HDL-C (mg/dL) decreased in two studies (p <0.01)(26,27), the same case for LDL-C (p < 0.05)(28). Also, azotemia, the CaxP(25), mean urinary indican, β2-microglobulin, mean C-reactive protein (CPR)(15), ionized calcium, albumin, alkaline phosphatase, and IL-6(26) decreased significantly at the end of the treatments, all with a p < 0.04. Otherwise, serum sodium(26), serum iron level, and mean transferrin saturation (TSAT)(15) increased, all present a p <0.005. The concentration of a relative abundance of *Bifidobacterium animalis* and *unclassified Blautia* (29) and mean fecal *Lactobacillales* and *Bifidobacteria* concentrations also increased(15).

### 3.2 Risk of bias in studies

Of the seven studies that underwent a risk of bias assessment and as a result 5 scored ‘some concerns’ and 2 ‘low’(26,28). The biases that were assessed were: selection (randomization sequence and concealment of allocation sequence), performance (masking of participants and personnel), detection (blinding of outcome assessors), and attrition (incomplete outcome data) and notification (selective notification of results).

All the studies analyzed were interventions, with the intention to treat, an excellent randomization of the patients in the control and intervention groups was shown in the seven studies analyzed. For this, software was used to select the patients and assign them to each group. The item with the highest risk of bias was information on data missing from the trials.

### 3.3 Network Meta-Analysis (inmunonutrition and Uremic Toxins)

The network diagram for each of the uremic toxins in the CKD stage 3 and 5 patients for each intervention group is shown in Figure 2.

**Figure 2.**
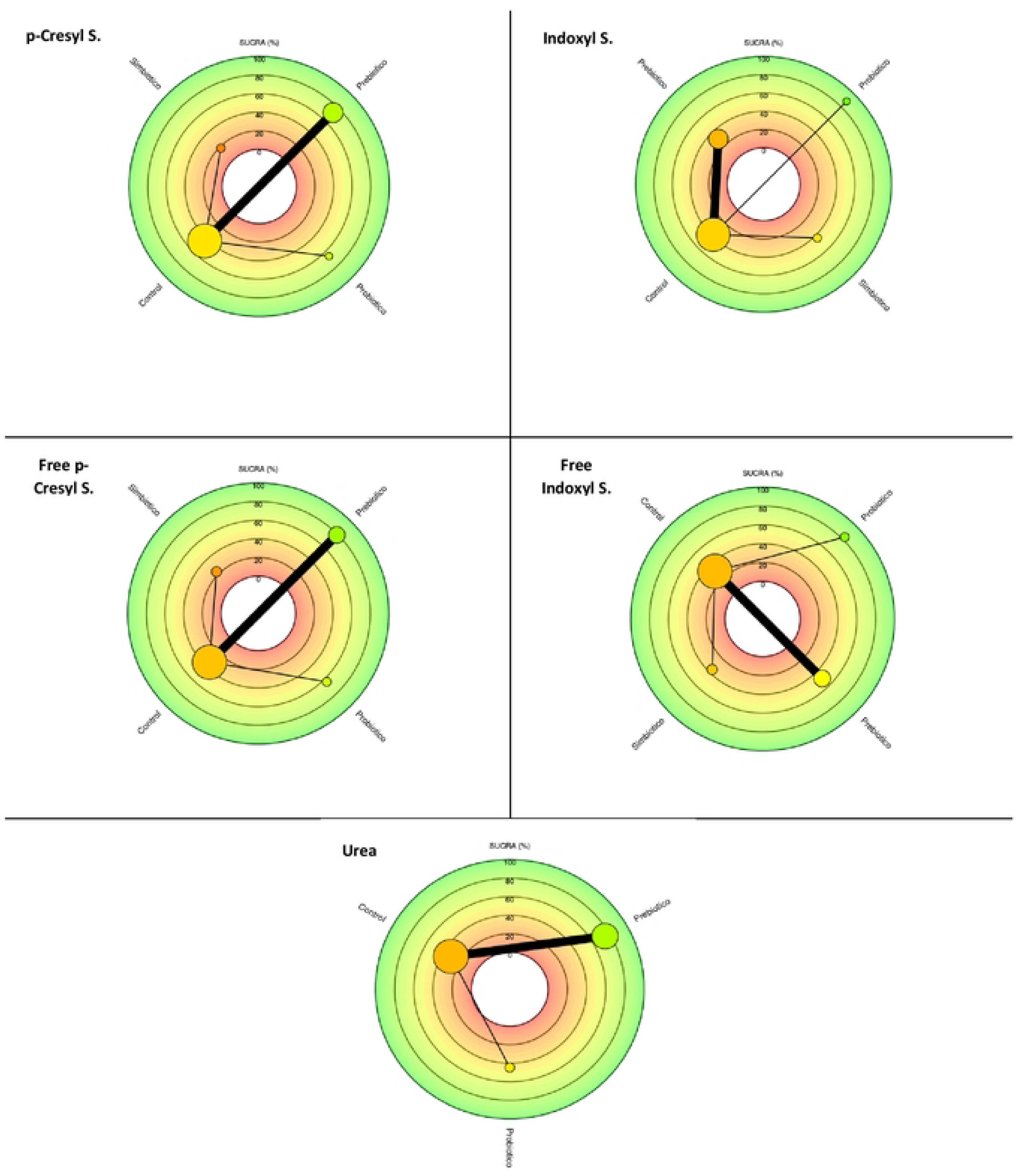
Network meta-analysis. Comparison of probiotics, prebiotics, symbiotic and control group In the reduction of uraemic toxins. The size of the nodes is proportional to the sample size of each immunonutrition intervention. Each uraemic toxin has a combination of interventions. For example, for the uraemic toxin p-Cresy1 Total, prebiotics (n=70), probiotics (n=24), symbiotics (n=28), as we.II as, the thickness of the lines is inversely proportional to the number of studies available. The number of studies for eachimmunonutrition intervention was as follows: Total p-Cresyl Sulfate (n=4), free p-Cresyl Sulfate (n=4), lndoxyl Sulfate (n=S); free indoxyl sulfate (n=S); Urea (n=S). About the type of intervention, 3 articles with the use of prebiotics, 2 with theuse of probiotics, and 2 with theuse of symbiotics are shown.

Table 2 shows the summary of the comparisons of the SUCRA values, which provides a relative classification of Immunonutrition. (Probiotics, prebiotics, and symbiotics) for each of the uremic toxins. In relation to the plots of the area under the curve for each uremic toxin are found in the supplementary material 1 F 1.

**Table 2.**
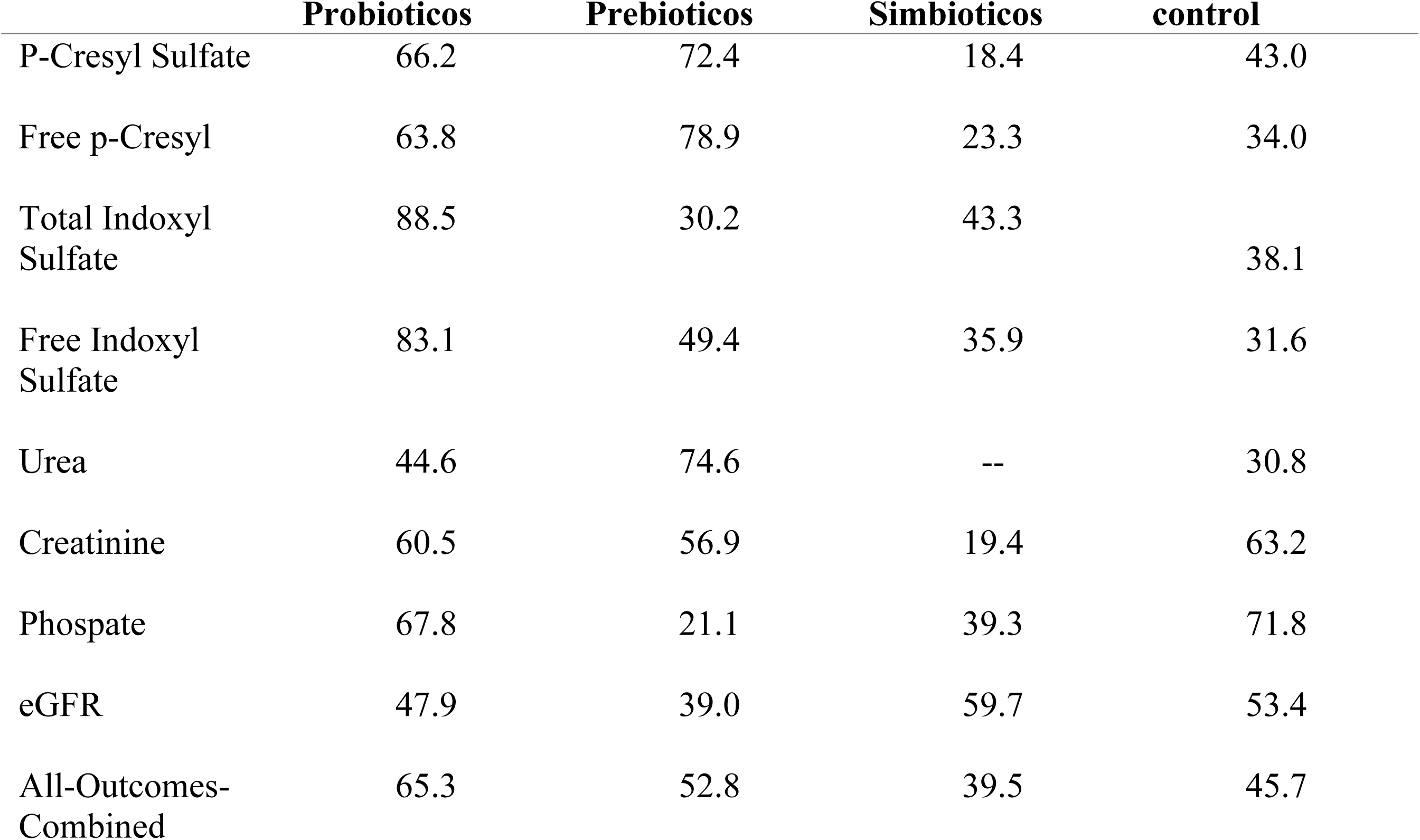
SUCRA values for each uremic toxin, by category combined.

|  | **Probiotics** | **Prebiotics** | **Simbiotics** | **control** |
| --- | --- | --- | --- | --- |
| P-Cresyl Sulfate | 66.2 | 72.4 | 18.4 | 43.0 |
| Free p-Cresyl | 63.8 | 78.9 | 23.3 | 34.0 |
| Total Indoxyl Sulfate | 88.5 | 30.2 | 43.3 | 38.1 |
| Free Indoxyl Sulfate | 83.1 | 49.4 | 35.9 | 31.6 |
| Urea | 44.6 | 74.6 | -- | 30.8 |
| Creatinine | 60.5 | 56.9 | 19.4 | 63.2 |
| Phospate | 67.8 | 21.1 | 39.3 | 71.8 |
| eGFR | 47.9 | 39.0 | 59.7 | 53.4 |
| All-Outcomes-Combined | 65.3 | 52.8 | 39.5 | 45.7 |

The use of prebiotics and probiotics was qualified mainly as the best option to reduce uraemic toxins, as is the case of p-Cresyl sulfate (SUCRA: 72.6%7 and 66.2 respectively). In relation to free p-Crasyl (SUCRA: 78.9% and 63.8% respectively), both indoxyl sulfate and free indoxyl sulfate are found only for the use of probiotics (SUCRA: 88.5% and 83.1%) finally, the use of prebiotics to decrease urea, cut with a SUCRA value; 74.6%.

There was no indication of potential effects in reducing uraemic toxins such as phosphate and creatinine, as well as benefiting the glomerular filtration rate (see supplementary material 2. F2).

### 3.4 Individual immunonutrition and reduction of uremic toxins

The implementation of prebiotics in the diet with interventions based on patients without renal function replacement therapy and stages 3 and 5 according to international guidelines. Said follow-up was carried out over a period of 12 months. A reduction in the accumulation of uremic toxins was found in these studies compared to placebos.

The treatment effect size in the PCS population was higher for probiotics (−8.95 95% CI: −53.3, 35.0), although this reduction was not significant. Similarly, when comparing total Indoxil sulfate, a larger effect size was observed for the probiotic group, however, the reduction was not significant (−6.39 95% CI: −16.0, 3.75). (Data are shown in supplementary material F3).

## 4. DISCUSSION

Preliminary evidence suggests that the modulation of the intestinal microbiota of CKD patients by oral supplementation with pre/probiotics or their combination (synbiotics), may decrease uremic toxins and inflammation, consequently improving endothelial function(29,32).

In renal replacement therapies, specifically in hemodialysis, Liu et al (33) demonstrated that the consumption of probiotics did not significantly alter species diversity of the fecal microbiome, however, it restores the community composition, with particular significance in non-diabetic HD patients (P = 0.007); specifically, probiotics raised the proportions of family *Bacteroidaceae* and *Enterococcaceae*, and reduced *Ruminococcaceae*, *Halomonadaceae*, *Peptostreptococcaceae*, *Clostridiales* Family XIII. Incertae Sedis and *Erysipelotrichaceae* in non-diabetic HD patients. Additionally, probiotics reduced the abundances of several uremic retention solutes in serum or feces, including indole-3-acetic Acid-O-glucuronide, 3-guanidinopropionic acid, and 1-methylinosine (P < 0.05). Also in HD therapy, Cruz-Mora et al (34) showed that symbiotic supplementation for 2 months significantly modifies the intestinal microbiota by increasing Bifidobacteria counts in patients (P < .0344).

On the other hand, in peritoneal dialysis therapy Pan et al (35) exposes that probiotics could significantly decrease the serum levels of high-sensitivity C-reactive protein and interleukin-6, which may indicate that both renal patients (who are not undergoing replacement therapy and those who are), are favored by the consumption of probiotics.

However, there are studies in which the symbiotic supplementation did not lower indoxyl sulphate toxin levels in comparation with the placebo group (p=0.438), neverthless it had a major effect in improving constipation and quality of life affected by constipation in patients undergoing chronic hemodialysis (36).

As is also the case with McFarlane et al (29) in which they present inconclusive results, since no differences were observed between free and total uremic toxins between placebo and symbiotic groups in a population in stage 3–4 chronic kidney disease.

Interestingly, patients assuming probiotics displayed a trend in reduction of the progression to end stage renal disease and dialysis initiation, in comparison with subjects assuming placebo (cumulative survival 57% vs. 34%, log rank p = 0.08, (Figure 2). No differences between the two groups were, however, observed in respect to the incidence of cardiovascular events (cumulative survival 81% vs. 89%, log-rank p = 0.55)

Alterations of the gut microbiota at various phases of CKD were also the focus of current research. It is important to explore the relationship between gut microbiota and CKD and investigate the difference in gut microbiota between patients with CKD and healthy people, as well as the different microbiota that could aggravate disease progression. Therefore, adjusting the use of probiotics, prebiotics, and symbiotics according to the distribution characteristics of floras at different phases has become a crucial measure to delay the course of CKD to ESRD.

All studies, including the control group, used various types of intervention delivery. It is important to point out that the outcome effects in the studies are controversial in terms of microbiota-modulating agents on endothelial function. In addition, it is important to point out cardiovascular markers, inflammation, and the progression of chronic kidney disease.

### 4.1 Implications for clinical practice

The results from our study on modulating the gut microbiome in chronic kidney disease (CKD) patients have significant implications for clinical practice. The findings suggest that interventions using probiotics, prebiotics, and synbiotics can effectively reduce uremic toxins such as indoxyl sulfate and p-cresyl sulfate, which are known to exacerbate CKD and its complications.

Incorporating these microbiome-modulating interventions could become a valuable adjunct therapy for CKD management. By reducing uremic toxins, these interventions may help mitigate inflammation and oxidative stress, potentially slowing the progression of CKD and improving patient outcomes. This approach could be particularly beneficial for patients in stages 3 to 5 of CKD, where traditional therapies alone may not sufficiently address the accumulation of these toxins.

Furthermore, synbiotics, which combine probiotics and prebiotics, appear to offer the most significant benefits, suggesting that a combined approach may be more effective than using either component alone. This insight could guide healthcare providers in designing more effective dietary and therapeutic strategies for CKD patients.

### 4.2 Study limitations

Regarding the limitations found in the studies analyzed, the limitation in the sample size must be noted, which must be considered to interpret the results, since it increases the risk of type I error. It would be convenient to carry out studies of these characteristics, metacentric, and with larger sample sizes. In addition, it would provide added value to include patients in other stages of the disease, as well as from the detection of the diagnosis. This would increase the heterogeneity of the population studied.

## 5. CONCLUSION

Studies report that a change in the composition of the intestinal microbiota of patients has a positive effect on the reduction of p-Cresyl and indoxyl sulfate, however, these changes are not consistent, and more randomized clinical trials with sample size are required. Larger studies confirm the effect of these different therapies on the modulation of the microbiota on intestinal bacterial composition and reductions in uremic toxins, with a broader detection than those already mentioned.

## Data Availability

All relevant data are within the manuscript and its Supporting Information files

## Acknowledgments

Appreciation to Susan Drier for her editing and style assessment.

## Authorship contributions

Study design: **MCB;** Data collection: **RCF;** Data analysis: **ICR;** Study supervision: **RP, MA;** Manuscript writing: **RCF, MCB;** Critical revisions for important intellectual content: **MK, RP**

## REFERENCES

1. Kotanko P, Carter M, Levin NW. Intestinal bacterial microflora-a potential source of chronic inflammation in patients with chronic kidney disease. Nephrol Dial Transpl [Internet]. 2006 [cited 2023 Mar 22];21:2057–60. Available from: https://academic.oup.com/ndt/article/21/8/2057/1820863

2. Pivari F, Mingione A, Piazzini G, Ceccarani C, Ottaviano E, Brasacchio C, et al. Curcumin Supplementation (Meriva®) Modulates Inflammation, Lipid Peroxidation and Gut Microbiota Composition in Chronic Kidney Disease. Nutrients. 2022;14(1).

3. Filipska I, Winiarska A, Knysak M, Stompór T. Contribution of Gut Microbiota-Derived Uremic Toxins to the Cardiovascular System Mineralization. Toxins (Basel). 2021 Apr;13(4).

4. Poesen R, Evenepoel P, De Loor H, Delcour JA, Courtin CM, Kuypers D, et al. The influence of prebiotic arabinoxylan oligosaccharides on microbiota derived uremic retention solutes in patients with chronic kidney disease: A randomized controlled trial. PLoS One. 2016 Apr 1;11(4).

5. Pandey KR, Naik SR, Vakil B V. Probiotics, prebiotics and synbiotics-a review. J Food Sci Technol. 2015 Dec;52(12):7577.

6. Chaudhari A, Dwivedi MK. The concept of probiotics, prebiotics, postbiotics, synbiotics, nutribiotics, and pharmabiotics. Probiotics Prev Manag Hum Dis. 2022 Jan;1–11.

7. Wang J, Cassone M, Gibson K, Lansing B, Mody L, Snitkin ES, et al. Gut Microbiota Features on Nursing Home Admission Are Associated With Subsequent Acquisition of Antibiotic-resistant Organism Colonization. Clin Infect Dis an Off Publ Infect Dis Soc Am. 2020 Dec;71(12):3244–7.

8. González Cordero EM, Cuevas-Budhart MA, Pérez Morán D, Trejo Villeda MA, Gomez-del-Pulgar G^a^-Madrid M. Relationship Between the Gut Microbiota and Alzheimer’s Disease: A Systematic Review. J Alzheimers Dis [Internet]. 2022 Mar 29 [cited 2022 Jun 21];87(2):519–28. Available from: https://pubmed.ncbi.nlm.nih.gov/35367961/

9. Wu D, Lewis ED, Pae M, Meydani SN. Nutritional Modulation of Immune Function: Analysis of Evidence, Mechanisms, and Clinical Relevance. Front Immunol [Internet]. 2019 [cited 2023 Aug 23];9(JAN). Available from: https://pubmed.ncbi.nlm.nih.gov/30697214/

10. Zapatera B, Prados A, Gómez-Martínez S, Marcos A. Immunonutrition: methodology and applications. Nutr Hosp. 2015;31:145–54.

11. Jovanovich A, Isakova T, Stubbs J. Microbiome and Cardiovascular Disease in CKD. Clin J Am Soc Nephrol. 2018 Oct;13(10):1598–604.

12. Hatem-Vaquero M, de Frutos S, Luengo A, González Abajo A, Griera M, Rodríguez-Puyol M, et al. Contribución de las toxinas urémicas a la fibrosis vascular asociada a la enfermedad renal crónica. Nefrología. 2018 Nov;38(6):639–46.

13. Cigarran Guldris S, González Parra E, Cases Amenós A. Microbiota intestinal en la enfermedad renal crónica. Nefrologia. 2017 Jan;37(1):9–19.

14. He S, Xiong Q, Tian C, Li L, Zhao J, Lin X, et al. Inulin-type prebiotics reduce serum uric acid levels via gut microbiota modulation: a randomized, controlled crossover trial in peritoneal dialysis patients. Eur J Nutr. 2022;61(2):665–77.

15. Simeoni M, Citraro ML, Cerantonio A, Deodato F, Provenzano M, Cianfrone P, et al. An open-label, randomized, placebo-controlled study on the effectiveness of a novel probiotics administration protocol (ProbiotiCKD) in patients with mild renal insufficiency (stage 3a of CKD). Eur J Nutr. 2019 Aug;58(5):2145–56.

16. Page MJ, Moher D, Bossuyt PM, Boutron I, Hoffmann TC, Mulrow CD, et al. PRISMA 2020 explanation and elaboration: updated guidance and exemplars for reporting systematic reviews. BMJ. 2021 Mar;372:n160.

17. Page MJ, McKenzie JE, Bossuyt PM, Boutron I, Hoffmann TC, Mulrow CD, et al. The PRISMA 2020 statement: an updated guideline for reporting systematic reviews. BMJ [Internet]. 2021 Mar 29 [cited 2022 Jul 25];372. Available from: https://www.bmj.com/content/372/bmj.n71

18. Olivo SA, Macedo LG, Gadotti C, Fuentes J, Stanton T, Magee DJ. Scales to Assess the Quality of Randomized Controlled Trials: A Systematic Review [Internet]. Vol. 156, Physical Therapy. 2008 [cited 2019 Jan 28]. Available from: www.ptjournal.org

19. Sterne JAC, Savović J, Page MJ, Elbers RG, Blencowe NS, Boutron I, et al. RoB 2: a revised tool for assessing risk of bias in randomised trials. 2023 [cited 2023 Mar 22]; Available from: 10.1136/bmj.l4898 http://www.bmj.com/

20. Higgins JPT, Altman DG, Gøtzsche PC, Jüni P, Moher D, Oxman AD, et al. The Cochrane Collaboration’s tool for assessing risk of bias in randomised trials. BMJ [Internet]. 2011 Oct 18 [cited 2023 Mar 22];343(7829). Available from: https://www.bmj.com/content/343/bmj.d5928

21. Jackson D, Barrett JK, Rice S, White IR, Higgins JPT. A design-by-treatment interaction model for network meta-analysis with random inconsistency effects. Stat Med. 2014;33(21):3639–54.

22. Higgins JPT, Jackson D, Barrett JK, Lu G, Ades AE, White IR. Consistency and inconsistency in network meta-analysis: concepts and models for multi-arm studies. Res Synth Methods. 2012;3(2):98–110.

23. Salanti G, Ades AE, Ioannidis JPA. Graphical methods and numerical summaries for presenting results from multiple-treatment meta-analysis: An overview and tutorial. J Clin Epidemiol [Internet]. 2011;64(2):163–71. Available from: 10.1016/j.jclinepi.2010.03.016

24. De Mauri A, Carrera D, Bagnati M, Rolla R, Chiarinotti D, Pane M, et al. Probiotics-Supplemented Low-Protein Diet for Microbiota Modulation in Patients with Advanced Chronic Kidney Disease (ProLowCKD): Results from a Placebo-Controlled Randomized Trial. Nutr 2022, Vol 14, Page 1637. 2022 Apr;14(8):1637.

25. Cosola C, Rocchetti MT, di Bari I, Acquaviva PM, Maranzano V, Corciulo S, et al. An Innovative Synbiotic Formulation Decreases Free Serum Indoxyl Sulfate, Small Intestine Permeability and Ameliorates Gastrointestinal Symptoms in a Randomized Pilot Trial in Stage IIIb-IV CKD Patients. Toxins (Basel). 2021 May;13(5).

26. Armani RG, Carvalho AB, Ramos CI, Hong V, Bortolotto LA, Cassiolato JL, et al. Effect of fructooligosaccharide on endothelial function in CKD patients: A randomized controlled trial. Nephrol Dial Transplant. 2022;37(1):85–91.

27. Ramos CI, Armani RG, Fernandes Canziani ME, Dalboni MA, Juliana C, Dolenga R, et al. Effect of prebiotic (fructooligosaccharide) on uremic toxins of chronic kidney disease patients: a randomized controlled trial.

28. Ebrahim Z, Proost S, Tito RY, Raes J, Glorieux G, Moosa MR, et al. The Effect of ß-Glucan Prebiotic on Kidney Function, Uremic Toxins and Gut Microbiome in Stage 3 to 5 Chronic Kidney Disease (CKD) Predialysis Participants: A Randomized Controlled Trial. Nutrients. 2022 Feb;14(4).

29. McFarlane C, Krishnasamy R, Stanton T, Savill E, Snelson M, Mihala G, et al. Synbiotics Easing Renal Failure by Improving Gut Microbiology II (SYNERGY II): A Feasibility Randomized Controlled Trial. Nutrients. 2021 Dec;13(12).

30. Cosola C, Rocchetti MT, Di Bari I, Acquaviva PM, Maranzano V, Corciulo S, et al. An innovative synbiotic formulation decreases free serum indoxyl sulfate, small intestine permeability and ameliorates gastrointestinal symptoms in a randomized pilot trial in stage IIIb-IV CKD patients. Toxins (Basel). 2021;13(5).

31. De Mauri A, Carrera D, Bagnati M, Rolla R, Vidali M, Chiarinotti D, et al. Probiotics-Supplemented Low-Protein Diet for Microbiota Modulation in Patients with Advanced Chronic Kidney Disease (ProLowCKD): Results from a Placebo-Controlled Randomized Trial. Nutrients. 2022 Apr;14(8).

32. Cosola C, Rocchetti MT, Cupisti A, Gesualdo L. Microbiota metabolites: Pivotal players of cardiovascular damage in chronic kidney disease. Pharmacol Res [Internet]. 2018;130(March):132–42. Available from: 10.1016/j.phrs.2018.03.003

33. Liu S, Liu H, Chen L, Liang SS, Shi K, Meng W, et al. Effect of probiotics on the intestinal microbiota of hemodialysis patients: a randomized trial. Eur J Nutr. 2020;59(8):3755–66.

34. Cruz-Mora J, Martínez-Hernández NE, Martín del Campo-López F, Viramontes-Hörner D, Vizmanos-Lamotte B, Muñoz-Valle JF, et al. Effects of a Symbiotic on Gut Microbiota in Mexican Patients With End-Stage Renal Disease. J Ren Nutr. 2014;24(5):330–5.

35. Pan Y, Yang L, Dai B, Lin B, Lin S, Lin E. Effects of Probiotics on Malnutrition and Health-Related Quality of Life in Patients Undergoing Peritoneal Dialysis: A Randomized Controlled Trial. J Ren Nutr. 2021;31(2):199–205.

36. Lydia A, Indra TA, Rizka A, Abdullah M. The effects of synbiotics on indoxyl sulphate level, constipation, and quality of life associated with constipation in chronic haemodialysis patients: a randomized controlled trial. BMC Nephrol. 2022 Dec;23(1):1–9.

37. Simeoni M, Citraro ML, Cerantonio A, Deodato F, Provenzano M, Cianfrone P, et al. An open-label, randomized, placebo-controlled study on the effectiveness of a novel probiotics administration protocol (ProbiotiCKD) in patients with mild renal insufficiency (stage 3a of CKD). Eur J Nutr. 2019 Aug;58(5):2145–56.

